# *ACE2* expression in adipose tissue is associated with COVID-19 cardio-metabolic risk factors and cell type composition

**DOI:** 10.1101/2020.08.11.20171108

**Authors:** Julia S. El-Sayed Moustafa, Anne U. Jackson, Sarah M. Brotman, Li Guan, Sergio Villicaña, Amy L. Roberts, Antonino Zito, Lori Bonnycastle, Michael R. Erdos, Narisu Narisu, Heather M. Stringham, Ryan Welch, Tingfen Yan, Timo Lakka, Stephen Parker, Jaakko Tuomilehto, Francis S. Collins, Päivi Pajukanta, Michael Boehnke, Heikki A. Koistinen, Markku Laakso, Mario Falchi, Jordana T. Bell, Laura J. Scott, Karen L. Mohlke, Kerrin S. Small

**Author notes:** These authors contributed equally to this work (Julia S. El-Sayed Moustafa, Anne U. Jackson, Sarah M. Brotman, Li Guan). Joint senior authors (Laura J. Scott, Karen L. Mohlke, Kerrin S. Small). Corresponding authors: Julia S. El-Sayed Moustafa Department of Twin Research and Genetic Epidemiology, King’s College London, London, UK, Kerrin S. Small, Department of Twin Research and Genetic Epidemiology, King’s College London, London, UK.

## Abstract

COVID-19 severity has varied widely, with demographic and cardio-metabolic factors increasing risk of severe reactions to SARS-CoV-2 infection, but the underlying mechanisms for this remain uncertain. We investigated phenotypic and genetic factors associated with subcutaneous adipose tissue expression of Angiotensin I Converting Enzyme 2 *(ACE2)*, which has been shown to act as a receptor for SARS-CoV-2 cellular entry. In a meta-analysis of three independent studies including up to 1,471 participants, lower adipose tissue *ACE2* expression was associated with adverse cardio-metabolic health indices including type 2 diabetes (T2D) and obesity status, higher serum fasting insulin and BMI, and lower serum HDL levels (*P*<5.32×10^-4^). *ACE2* expression levels were also associated with estimated proportions of cell types in adipose tissue; lower *ACE2* expression was associated with a lower proportion of microvascular endothelial cells (*P*=4.25×10^-4^) and higher macrophage proportion (*P*=2.74×10^-5^), suggesting a link to inflammation. Despite an estimated heritability of 32%, we did not identify any proximal or distal genetic variants (eQTLs) associated with adipose tissue *ACE2* expression. Our results demonstrate that at-risk individuals have lower background *ACE2* levels in this highly relevant tissue. Further studies will be required to establish how this may contribute to increased COVID-19 severity.

## Main

Since December 2019 the COVID-19 pandemic has swept across the world, with over sixteen million confirmed cases worldwide and almost 700,000 fatalities reported at the time of writing^1^. A distinctive and challenging aspect of COVID-19 is its wide disease spectrum, with vast differences observed in both symptom range and severity, as well as clinical progression, across those affected^2^. While initial reports focused on respiratory symptoms^3,4^, it is now clear that severe COVID-19 also has a substantial effect on the cardiovascular and renal systems^5^. Marked differences in clinical outcomes have been associated with demographic features including age and sex, as well as comorbidities including cardiovascular disease, diabetes, and obesity^2,4,5^. An urgent need exists to improve understanding of the mechanisms by which such predisposing factors increase susceptibility to severe reactions to SARS-CoV-2 infection.

ACE2 has become central in understanding COVID-19 pathogenesis, as SARS-CoV-2 employs ACE2 as a receptor for cellular entry^6,7^. *ACE2* is expressed in a number of tissues and cell types^6,8-11^. It is currently unclear whether higher ACE2 levels are harmful or beneficial in the context of COVID-19. Higher levels of ACE2 may on the one hand provide additional targets enabling viral invasion of *ACE2*-expressing cells, however higher ACE2 also has beneficial effects in regulation of the renin-angiotensin system (RAS), which is critical for controlling hypertension and associated cardio-metabolic disorders^12,13^. Extensive reports of adverse cardio-metabolic effects in rodent models with decreased or ablated ACE2 levels support the critical role of ACE2 in a healthy cardiovascular system^14^.

Obesity, characterised by an excess of fat mass^15^, is one of the strongest reported risk factors for severe COVID-19^2,5^, but the underlying mechanism of this increased risk remains unclear. Adipose tissue, composed of adipocytes, immune and vascular endothelial cells, is both an energy reservoir and an endocrine organ, and secretes a wide variety of hormones, cytokines and chemokines^16,17^. In the Genotype-Tissue Expression (GTEx) resource of gene expression in 54 tissues, adipose tissue ranks among the highest sites of *ACE2* expression in the body^11,18^ (Supplementary Figure 1). Several potential mechanisms have been proposed for the contribution of adipose tissue to COVID-19 clinical severity^19,20^, and its influence may in fact be multi-factorial. Adipose tissue has been shown to act as a viral reservoir for numerous viruses, including SARS-CoV and type A influenza^21,22^, and may contribute to prolonged viral shedding^23^. Critically, *ACE2* in adipose tissue plays an important role in balancing of the local adipose RAS, disruption of which may then lead to wider systemic effects on the renin-angiotensin system^20^. Adipose tissue is also a prime candidate contributor to the widely-reported cytokine storm characteristic of severe COVID-19^19,20^. Most importantly, obesity is a potentially modifiable COVID-19 risk factor, and deeper understanding of the contribution of adipose tissue to COVID-19 severity, as well as to cardio-metabolic risk more generally, may better inform therapeutic strategies, and motivate policies supporting healthy weight maintenance programmes.

Given the dual relevance of ACE2 in maintenance of cardio-metabolic health generally as well as its role as SARS-CoV-2 receptor, it is of interest to explore whether *ACE2* expression is associated with demographic and phenotypic traits associated with COVID-19 severity. There have been several reports supporting variation in *ACE2* expression across tissues, and early evidence of differences in associations across samples and even between cell types within the same tissue^11,24^.In this study, we investigated the phenotypic and genetic factors associated with *ACE2* gene expression in subcutaneous adipose tissue from 1,471 participants from three cohorts^25-27^, representing the largest meta-analysis of adipose tissue gene expression studies to date. We quantified adipose *ACE2* expression levels in bulk RNA-Seq data from TwinsUK^25,28,29^ (765 female mono- and dizygotic twins), 426 males from the Metabolic Syndrome in Men (METSIM) study^26^, and 149 Finnish males and 131 Finnish females from the Finland-United States Investigation of NIDDM Genetics (FUSION) Tissue Biopsy Study^27^. The cohorts differ in country of origin, male:female composition, age distribution and underlying ascertainment criteria (Table 1). Study participants ranged in age from 35 to 85 years, with variation between studies (Table 1 and Supplementary Figure 2). While TwinsUK and METSIM are representative of the UK and Kuopio Finland populations, the FUSION study design was enriched for individuals with abnormal glycaemic indices at time of biopsy.

**Figure 1:**
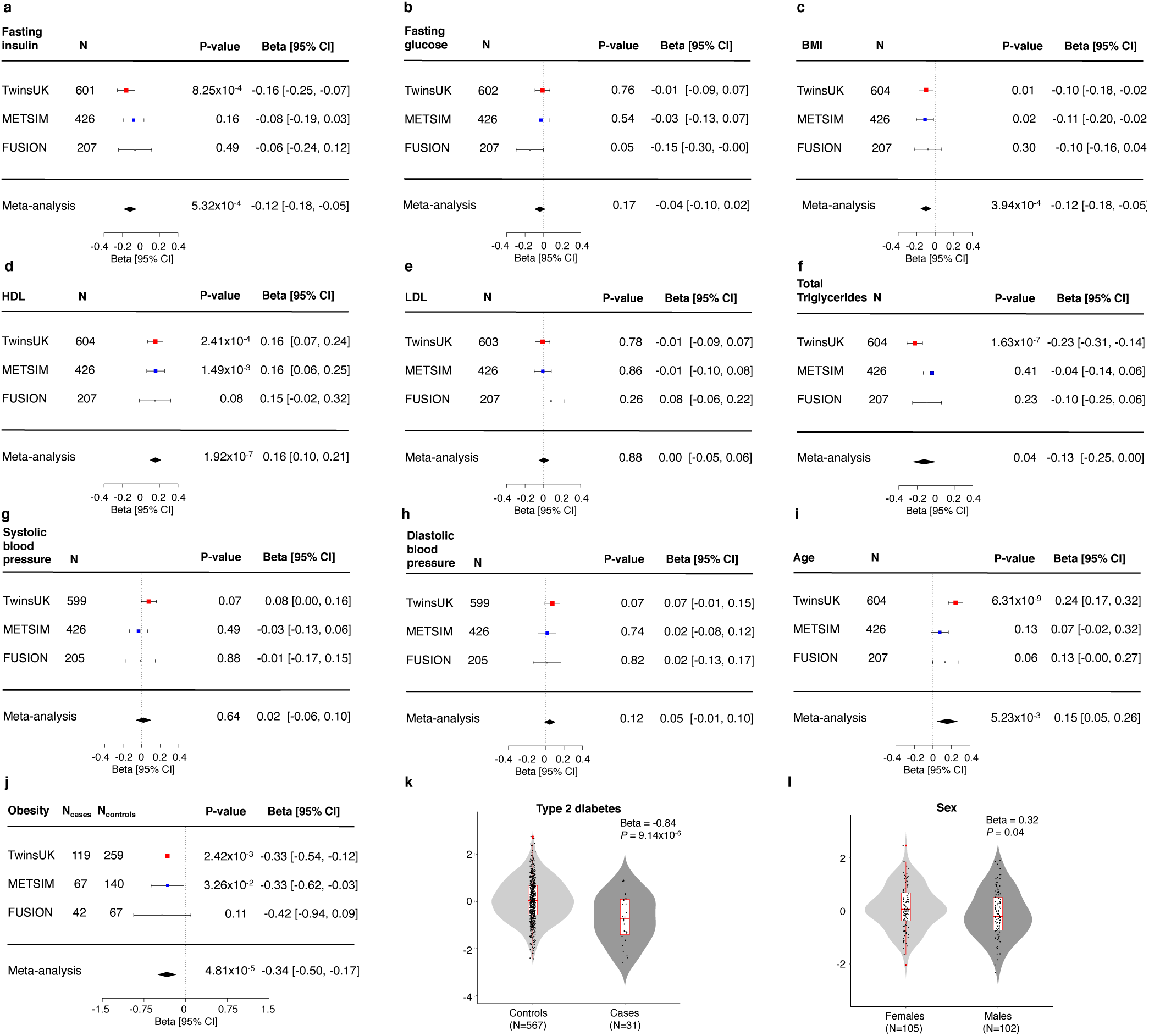
Associations of adipose tissue *ACE2* expression levels with cardio-metabolic and demographic traits in the TwinsUK, METSIM and FUSION studies. Squares with error bars represent the standardised β coefficients and their 95% confidence intervals for association of *ACE2* expression levels with each of the traits (derived from linear/linear mixed effects regression models), with the meta-analysis effect sizes and 95% confidence intervals (random-effects meta-analysis) shown as black diamonds. N represents the sample size for each analysis. a) serum fasting insulin, b) fasting plasma glucose, c) BMI d) serum HDL cholesterol, e) serum LDL cholesterol, f) serum total triglycerides, g) systolic blood pressure, h) diastolic blood pressure, i) age, j) obesity status. Association of *ACE2* expression with k) type 2 diabetes status in TwinsUK and l) sex in the FUSION study. Boxplots (k-l) display the median and inter-quartile range (IQR), with whiskers corresponding to +/- 1.5*IQR.

**Figure 2:**
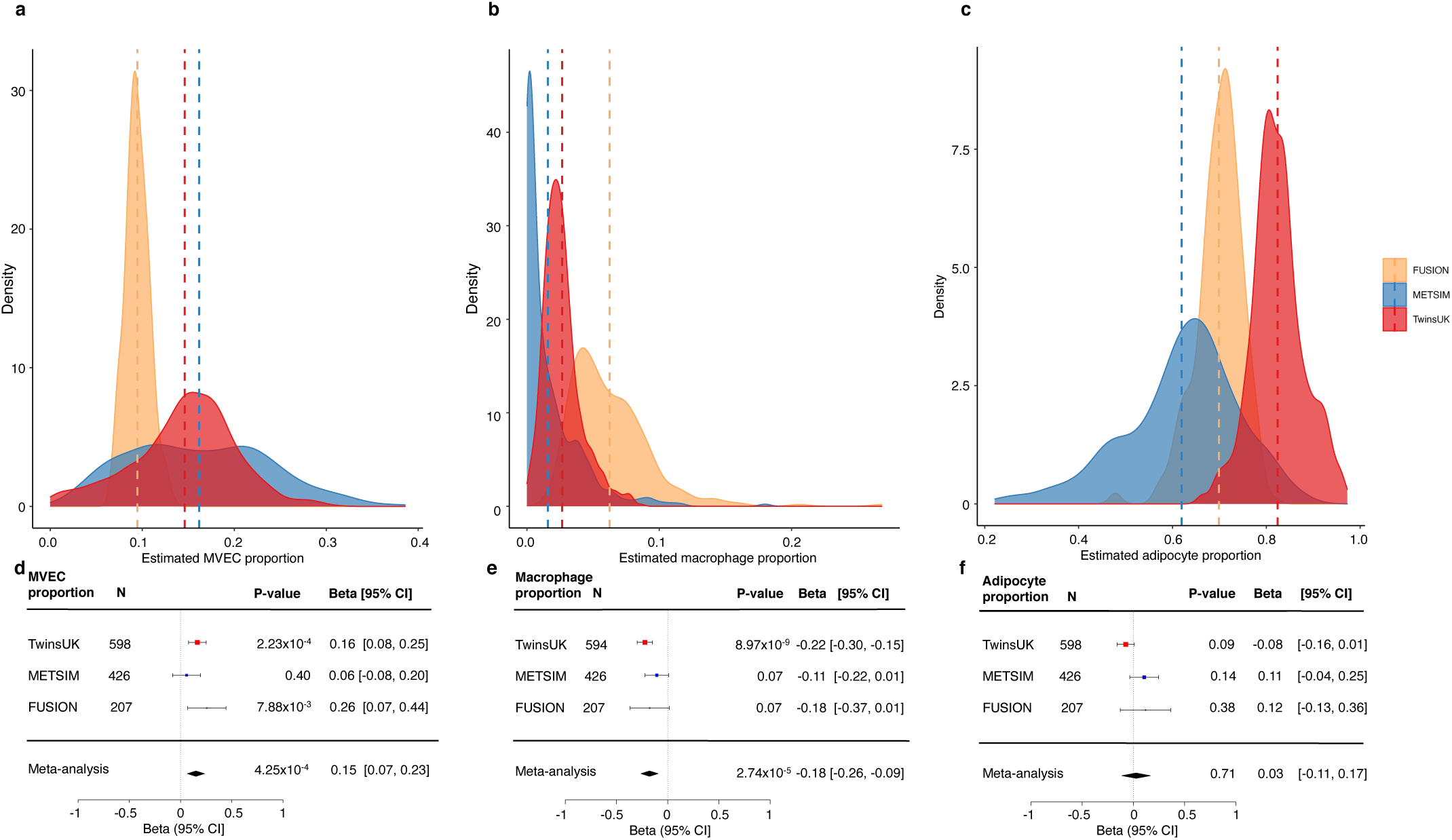
Adipose tissue estimated cell type proportions and their association with *ACE2* expression levels across the TwinsUK, METSIM and FUSION studies. Density plots of estimated cell type proportions of a) microvascular endothelial cells (MVEC), b) macrophages, c) adipocytes in each of the TwinsUK, METSIM and FUSION studies. Vertical dashed lines correspond to the study mean. d-f) Association of adipose tissue *ACE2* expression levels with estimated d) MVEC, e) macrophage and f) adipocyte proportions. Squares with error bars represent the standardised β coefficients and their 95% confidence intervals for association of *ACE2* expression levels with each of the traits (derived from linear/linear mixed effects regression models), with the meta-analysis effect sizes and 95% confidence intervals (random effects meta-analysis) shown as black diamonds. N represents the sample size for each analysis.

**Table 1:** Descriptive statistics of study participants. Quantitative traits are reported as median[1^st^-3^rd^ quartiles]. Statistics for all traits are calculated including only non-diabetic participants. *ACE2* expression levels are reported as TMM-adjusted counts per million (CPMs).

|  | **TwinsUK** | **METSIM** | **FUSION** |
| --- | --- | --- | --- |
| **N [% female]** | 765[100%] | 426[0%] | 280[47%] |
| **Age (years)** | 59[52-65] | 54 [51-59] | 60[55-65] |
| **BMI (kg/m<sup>2</sup>)** | 25.6[23.3-28.8] | 26.2 [24.6-28.6] | 26.7[24.2-28.8] |
| **Serum fasting insulin (pmol/l)** | 38.0[25.0-59.5] | 35.4 [25.2-55.8] | 43.8[28.8-59.1] |
| **Fasting plasma glucose (mmol/l)** | 4.9[4.6-5.2] | 5.7[5.4-6.0] | 5.9[5.6-6.3] |
| **Serum HDL (mmol/l)** | 1.80[1.53-2.14] | 1.45 [1.20-1.70] | 1.44[1.20-1.70] |
| **Serum LDL (mmol/l)** | 3.18[2.57-3.82] | 3.43 [2.96-3.96] | 3.34[2.78-3.89] |
| **Serum triglycerides (mmol/l)** | 0.96[0.72-1.33] | 1.17 [0.88-1.62] | 1.17[0.89-1.56] |
| **Systolic BP (mmHg)** | 128[118-139] | 132[123-141] | 133[121-145] |
| **Diastolic BP (mmHg)** | 78[71-85] | 87[81-93] | 82[76-87] |
| **Adipose tissue *ACE2* expression (TMM-adjusted CPMs)** | 1.86[1.16-3.05] | 1.25[0.82-1.83] | 1.15[1.07-2.62] |

We hypothesised that the true effect sizes of *ACE2* gene expression-phenotypic associations may vary between cohorts differing in demographic characteristics, and due to technical and methodological differences between the studies. We therefore assessed association of *ACE2* gene expression with demographic and phenotypic traits in each of the TwinsUK, METSIM and FUSION studies, adjusting for age and BMI, and then conducted a random-effect inverse-variance-weighted meta-analysis combining association results from the three studies. Results of meta-analysis of the three studies showed consistency in association signals across cohorts for some traits, but heterogeneity for others.

Adipose tissue *ACE2* expression was relatively low across all three cohorts (Table 1, Supplementary Figures 3 and 4). In our meta-analysis, lower adipose tissue *ACE2* expression levels were associated with higher serum fasting insulin (β [95% CI] = - 0.12[-0.18, -0.05]; *P* = 5.32×10^-4^), and higher body mass index (BMI) (β [95% CI]= - 0. 10[-0.16, -0.04]; *P* = 3.94×10^-4^) (Figure 1 and Supplementary Table 1). Consistent with the association observed for BMI, obese participants had lower adipose tissue *ACE2* expression compared to normal-weight controls (β [95% CI]= -0.34[-0.50, -0.17]; *P* = 4.81×10^-5^) (Figure 1 and Supplementary Table 1).

TwinsUK included a subset of individuals diagnosed with T2D prior to biopsy collection. Within TwinsUK, T2D status was associated with lower adipose tissue *ACE2* expression compared to normoglycaemic controls (β [95% CI] = -0.84[-1.21, -0.47]; *P* = 9.14×10^-6^). No association was observed with fasting glucose in our study (P>0.05) (Figure 1 and Supplementary Table 1).

Lower *ACE2* expression was strongly associated with lower serum HDL cholesterol (β [95% CI] = 0.16[0.10, 0.21]; *P* = 1.92×10^-7^) (Figure 1 and Supplementary Table 1). Association of total serum triglycerides with adipose tissue *ACE2* expression showed heterogeneity between studies (I^2^=73.24; Q-test *P* = 0.01). Results of meta-analysis of all three studies showed only nominally-significant association between lower *ACE2* expression and higher triglyceride levels (β [95% CI] = -0.13[-0.25, 0.00]; *P* = 4.15×10^-2^), but this association was not significant after multiple testing correction (MTC threshold *P* < 3.33×10^-3^). In TwinsUK, lower *ACE2* expression in adipose tissue was strongly associated with higher total triglyceride levels (β [95% CI] = -0.23[-0.31, -0.14]; *P* = 1.63×10^-7^). Adipose tissue *ACE2* expression was not associated with serum LDL cholesterol, nor with systolic or diastolic blood pressure *(P* > 0.05) in our study (Figure 1 and Supplementary Table 1).

Severity of COVID-19 disease presentation has been associated with age^2-5^. Upon meta-analysis, higher adipose *ACE2* expression levels were nominally associated with older age (β [95% CI] = 0.15 [0.05, 0.26]; *P* = 5.23×10^-3^), but not significant after multiple testing correction (Figure 1 and Supplementary Table 1). However, we observed heterogeneity in age effects between studies (Heterogeneity I^2^=71.38; Cochrane’s Q-test *P* = 0.02) potentially driven by differences in the age distribution of studies. In the study with the largest age range, TwinsUK (38-85 years), higher adipose tissue *ACE2* expression was strongly associated with older age (β [95% CI] = 0.25[0.17, 0.32]; *P* = 6.31×10^-9^). Higher *ACE2* expression in older individuals observed in adipose tissue was consistent with previous reports of age-dependent *ACE2* expression in nasal epithelium^30^. However, in skin of the same TwinsUK participants lower *ACE2* expression was associated with increased age (β [95% CI]= -0.23[-0.31, -0.14]; *P* = 4.77×10^-7^), supporting heterogeneity of effects between tissues^11^ (Supplementary Figure 5).

Located on the X chromosome, *ACE2* has been shown to escape from X inactivation^31^. Association of *ACE2* expression with sex was assessed only in the FUSION study, which included both males and females. Consistent with reports from GTEx^11^, *ACE2* expression was nominally higher in females in FUSION (β [95% CI] = 0.32[0.02, 0.61]; *P* = 0.037), but this difference was not significant after multiple testing correction (Figure 1 and Supplementary Table 1).

Adipose tissue consists of multiple cell types, gene expression of all of which is represented in bulk RNA-Seq data, and to our knowledge, it is still unclear which adipose cell types are responsible for expression of *ACE2*. We therefore assessed the relationship between *ACE2* expression in subcutaneous adipose tissue and estimated adipose tissue cell type proportions. Consistent with the differences in biopsy methods employed, significant heterogeneity in cell type composition was observed between the three studies (Figure 2). Despite these differences, higher proportion of microvascular endothelial cells (MVEC) was associated with higher *ACE2* expression (β [95% CI]= 0.15[0.07, 0.23]; *P* = 4.25×10^-4^), suggesting microvascular endothelial cell subpopulations contribute to *ACE2* expression levels in adipose tissue (Figure 2 and Supplementary Table 2). Conversely, lower *ACE2* expression was associated with higher macrophage proportion (β [95% CI] = -0.18 [-0.26, -0.09]; *P* = 2.74×10^-5^) (Figure 2 and Supplementary Table 2). Increased macrophage infiltration in adipose tissue has been associated with a pro-inflammatory state^32^. *ACE2* expression was not associated with adipocyte proportion (*P* > 0.05; Figure 2 and Supplementary Table 2).

To establish whether phenotypic associations with *ACE2* expression were simply reflecting differences in adipose tissue cell type composition^33^, we conducted sensitivity analyses including the estimated proportion of MVECs as a covariate in our models. While strength of association was attenuated for some phenotypes, the associations were largely robust to adjustment for MVEC proportion (Supplementary Table 3), suggesting phenotypic associations were not driven solely by differences in the proportions of *ACE2*-expressing cell types. Furthermore, correlation patterns between phenotypic traits including age and each of adipose tissue *ACE2* expression and MVEC proportions were notably different (Supplementary Figure 6). In contrast, phenotypic associations were notably attenuated upon inclusion of both MVEC and macrophage proportions as covariates in our models (Supplementary Table 4), potentially due to adipose tissue macrophage proportions reflecting inflammatory status.

There is evidence that genetic factors may influence risk of severe COVID-19^34^. Using the twin structure of TwinsUK, we estimated the heritability of adipose tissue *ACE2* expression to be 0.32[95% CI = 0.14-0.50] (Figure 3). To identify proximal genetic variants associated with *ACE2* expression, we conducted a cis-eQTL meta-analysis of all three studies including 1,151 participants for whom chromosome X genotype data were available (See Methods section). No genetic variants within 1Mb of *ACE2* were associated with adipose tissue *ACE2* expression after correction for multiple testing (minimum *P* = 2.58×10^-3^ at chrX:15263126_A/C) (Figure 3 and Supplementary Table 5). This result was consistent with the absence of an *ACE2* cis-eQTL signal in numerous GTEx tissues^11^. We also assessed whether any trans-eQTL variants genome-wide were associated with *ACE2* expression levels; no variants were associated with *ACE2* expression at genome-wide significance (Figure 3 and Supplementary Table 6). Neither adjustment for MVEC proportion nor meta-analysis of males and females separately identified significant cis- or trans-eQTLs in our analyses.

**Figure 3:**
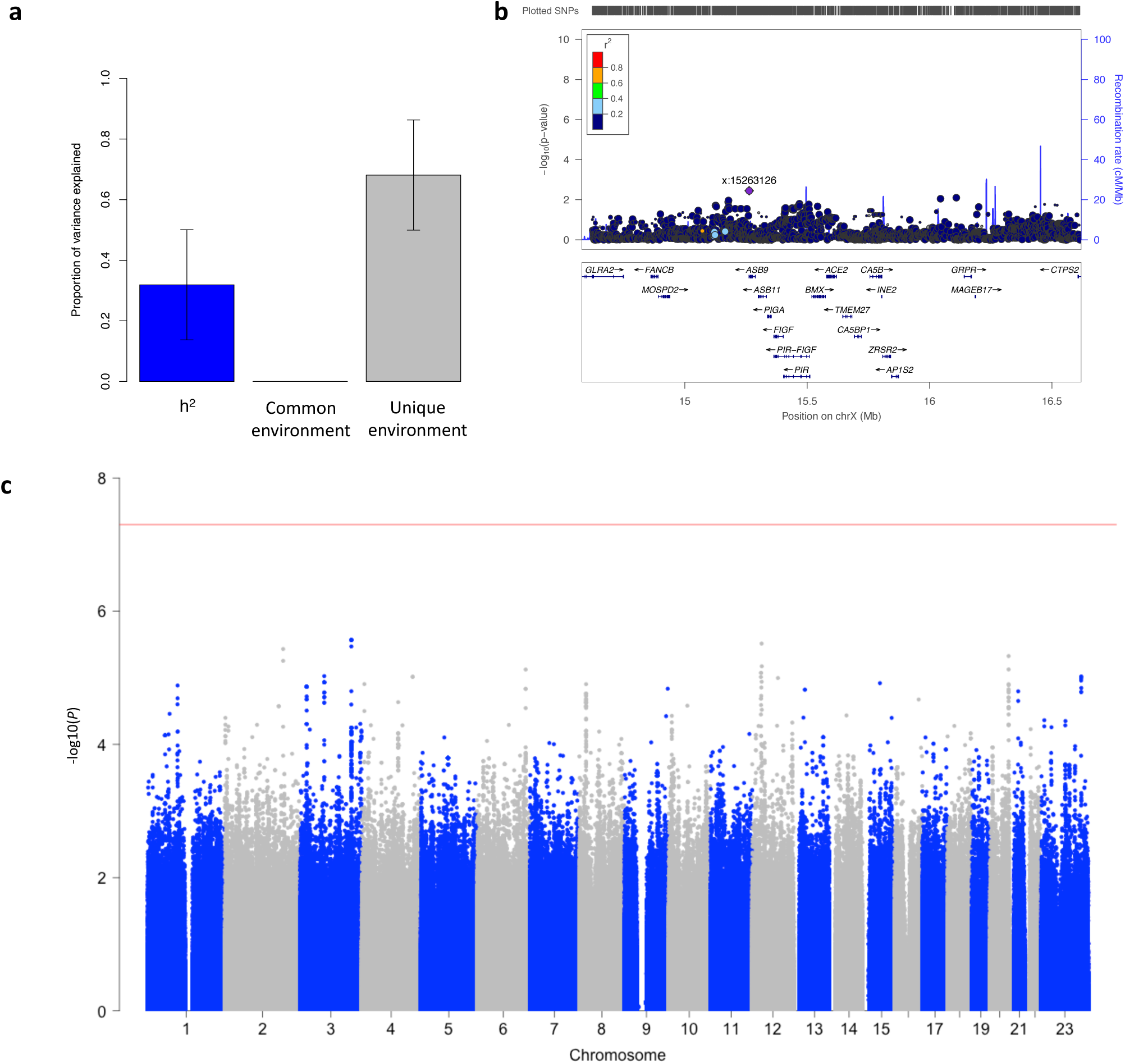
Genetic factors influencing adipose tissue expression of *ACE2*. a) Heritability of *ACE2* expression, assessed using an ACE model in the TwinsUK study. *ACE2* expression was partitioned into that which could be explained by (A) additive genetic effects (h^2^), common environment (C) and unique environment (E). Error bars show the 95% confidence intervals, b) Regional plot of *ACE2* cis-eQTL associations in 1,157 participants, within 1Mb either side of the *ACE2* transcription start site, c) Manhattan plot of genome-wide association study meta-analysis of adipose tissue *ACE2* expression in 1,428 participants. For plots b-c, the x-axis denotes chromosomal position, and the y-axis is -log10(P-value). The red line indicates the threshold for genome-wide significance (P < 5×10^-8^).

Finally, we explored epigenetic effects on *ACE2* expression using matched adipose tissue methylation data, measured on the Illumina Infinium HumanMethylation450 BeadChip for 540 participants in TwinsUK^35^. Similar to our eQTL results, no proximal methylation probes were associated with genetic variants in cis or trans. In addition, no proximal methylation probes were associated with adipose *ACE2* expression (minimum *P* = 0.048 at cg08559914), indicating that methylation at measured sites in our sample does not appear to be a reliable proxy for adipose tissue gene expression at this locus (Supplementary Figure 7).

Taken together, the analyses presented here show that *ACE2* expression in adipose tissue is associated with multiple COVID-19 comorbidities. We have shown that lower adipose tissue *ACE2* expression was associated with diabetes and obesity status, increased serum fasting insulin and triglyceride levels, BMI, and with increased macrophage infiltration in adipose tissue, a marker of inflammation^32^. *ACE2* expression was positively correlated with HDL levels, age and adipose tissue MVEC proportion. As previously noted for *ACE2* associations reported in other tissues^36^, given its association with adipose tissue cell type proportions, it remains unclear to what extent these associations are mediated through differences in underlying cell type composition. While sensitivity analyses adjusting for MVEC cell type proportion estimates suggested that the *ACE2* expression-trait associations were robust, estimation of cell type composition from bulk RNA-Seq data in non-blood tissues remains in its infancy. Adipose tissue single-cell RNA sequencing (scRNA-Seq) datasets will be critical to fully address this question. Furthermore, as seen for its association with age in the TwinsUK sample, *ACE2* expression-trait associations may differ between tissues, supporting the need for caution in their interpretation and extrapolation. The contrast between our observation that adipose tissue *ACE2* levels were positively correlated with age -another strong risk factor for severe COVID-19 yet negatively correlated with COVID-19 cardio-metabolic risk factors may reflect the multi-systemic nature of COVID-19, and may indicate that the various COVID-19 risk factors, including age, may be mediated through different tissues and underlying mechanisms of susceptibility. Despite evidence of heritability of adipose tissue *ACE2* expression levels and our meta-analysis sample size, we did not yet identify cis- or trans-eQTLs associated with adipose *ACE2* expression. Increased sample sizes may be needed to detect genetic factors influencing expression of *ACE2* in this important tissue.

Overall, our results suggest that lower adipose tissue expression of *ACE2* is associated with multiple adverse cardio-metabolic health indices, all of which are risk factors for severe COVID-19^2,4,5^. Increasing evidence suggests that COVID-19 severity may be modulated by widespread microvascular damage and increased predisposition to thrombotic events^37-39^, in which ACE2, and the RAS system more widely, may play a critical role. Adipose tissue is an active endocrine organ which secretes hormones, chemokines and cytokines that contribute to regulation of inflammatory and immune responses^16,17^. Given the association of *ACE2* expression with cardio-metabolic risk factors for severe COVID-19, it is intriguing to consider whether low levels of adipose *ACE2* expression may contribute to COVID-19 progression in at-risk individuals. SARS-CoV infection has been shown to reduce ACE2 levels post-infection^40-43^. Our observation that individuals suffering from recognised severe COVID-19 comorbidities may have lower pre-infection ACE2 levels in some tissues may support the theory that low ACE2 levels in at risk-individuals are then reduced further upon SARS-CoV-2 infection, contributing to the dysregulation of the RAS characteristic of severe COVID-19^44^. Further studies will be needed to clarify the contribution of adipose tissue *ACE2* expression levels to COVID-19 and beyond.

## Materials and methods

### Sample collection

#### TwinsUK

Gene expression data, measured by RNA-Seq, was available for up to 804 female twins from the TwinsUK cohort^45^. Sample collection and processing in TwinsUK has been reported fully elsewhere^45^. Briefly, punch biopsies were taken from a sun-sprotected area of the abdomen from each participant. Subcutaneous adipose tissue and skin were separated and RNA extracted for each tissue type. Lymphoblastoid cell lines (LCL) were generated from blood samples collected at the time of biopsy. Gene expression in adipose tissue (n=765), skin (n=706), LCLs (n=804) and whole blood (n=389) was measured by RNA sequencing, as previously described^45^.

#### METSIM

Sample collection and processing has previously been reported for the population-based METSIM cohort, composed of 10,197 males of Finnish ancestry^46^. RNA-sequencing of subcutaneous adipose tissue for 426 male participants collected near the umbilicus by needle biopsy was generated as previously described^26^.

#### FUSION

331 living FUSION participants were recruited from Helsinki, Savitaipale, and Kuopio, as previously described^27^. Adipose tissue biopsies were obtained and processed from 296 of the 331 participants following the same general protocol as previously described for the concurrently-obtained muscle biopsies^27^. Biopsies were taken under local anaesthetic, by surgical scalpel from abdominal subcutaneous fat 5-10 cm lateral of the umbilicus, cleaned of blood and other visible non-adipose tissue, and rinsed with 0.9% sodium chloride. Adipose RNA-sequencing was performed as described for muscle RNA-sequencing^27^.

### RNA-Seq data processing

#### TwinsUK and METSIM

RNA-Seq reads were aligned to the hg19 reference genome using STAR^47^ version 2.4.0.1 (TwinsUK) and 2.4.2a (METSIM), and quality control conducted as previously described^26,48^. Gene-level quantification was conducted using the quan function from QTLtools^49^ and Gencode version 19^50^.

#### FUSION

RNA-Seq reads were aligned to the hg19 reference genome using procedures as described in Scott *et al*^27^. Six outlier samples were excluded based on read coverage at the 3’ end of the gene body, and five outliers based on gene diversity. The allelic RNA-Seq read count distribution was compared to known sample genotypes using verifyBamID^51^ and two contaminated samples were identified and removed, as well as one pair of sample swaps corrected. Reported sex of the remaining samples was verified by matching the sex inferred using *XIST* gene expression and the mean Y chromosome gene expression. Linear regression of gene expression as a function of age, sex, batch, and RIN was performed in addition to performing PCA on the gene expression residuals. The minimum number of PCs were selected to explain 20% of the variance in gene expression. The PCs were transformed to z-scores and did not observe any sample outliers with |z-score| >5. Finally, one non-Finnish subject and one from each of two first-degree relative pairs were removed. A total of 280 samples were retained in the analysis.

### Gene inclusion and normalization

For all three studies, RNA-Seq gene expression data were filtered to retain only genes with 5 or more counts per million (CPMs) in greater than or equal to 25% of individuals in each study. Trimmed mean of M-values normalisation was applied to gene counts^52^, and TMM-normalised gene CPMs were then inverse-normalised prior to all downstream analyses (inverse-normalized *ACE2)*.

### Estimation of adipose tissue cell type proportions

Adipose tissue cell type proportions were estimated in TwinsUK using CIBERSORT^53^, as previously described^33^. Estimated cell types included in our analysis were adipocytes, microvascular endothelial cells (MVEC) and macrophages.

To estimate the proportions of adipose and blood cell types in the adipose biopsies from the FUSION and METSIM studies, a reference transcriptome was created using RNA-Seq of whole blood (GEO accession GSE67488^54^), and of four types of purified adipose cells (adipocytes, macrophages, CD4+ T cells, and microvascular endothelial cells) described in Glastonbury *et al*^33^. For the FUSION study, reference transcriptome reads were aligned to the hg19 reference genome using the same read mapping and quality control procedure as used for the FUSION adipose RNA-Seq data^27^. Blood/cell-type proportions for each FUSION adipose sample were estimated using the unmix function from DESeq2 v1.18.1^55^. A gene expression signature matrix was constructed and blood/cell-type proportions were estimated for each METSIM adipose sample using CIBERSORT^53^.

### Phenotypic association analyses

For quantitative phenotypic trait association analyses, in TwinsUK we excluded a subset of participants who had not fasted on the day of biopsy and for whom biochemical measurements were not matched to the day of biopsy, or for whom fasting and time of visit covariates were not available (n=161). Participants with T2D were excluded from all analyses in all studies except those modelling association of diabetes status with *ACE2* expression.

We assessed association between *ACE2* gene expression levels (Gencode ID ENSG00000130234.6) and phenotypic traits including age, BMI, serum fasting insulin, fasting plasma glucose, serum HDL and LDL cholesterol, total triglycerides, systolic and diastolic blood pressure, obesity, T2D status (TwinsUK only) and sex (FUSION only). For obesity status, cases (TwinsUK: n=119; METSIM: n=67; FUSION: n=68) were defined as participants with BMI ≥ 30kg/m^2^, while controls (TwinsUK: n=259; METSIM: n=140; FUSION: n=79) were defined as participants with BMI < 25kg/m^2^. In TwinsUK, established T2D case status (n=31) was defined through longitudinal patient self-reports, and longitudinal records of elevated fasting plasma glucose (≥ 7mmol/l) and serum fasting insulin levels. In this study, individuals were defined as T2D controls (n=567) if they had no longitudinal records of elevated fasting plasma glucose nor serum fasting insulin, as well as no self-reports of diabetes. Individuals with insulin resistance, defined as participants with longitudinally-elevated serum fasting insulin levels (>60pmol/l) with fasting plasma glucose levels consistently below 6.0mmol/l (n=130),were excluded from T2D association analyses. In FUSION, study participants underwent a 2-h, four-point OGTT after a 12-h overnight fast, and T2D, IFG, IGT and NGT status was assigned per WHO criteria^56^. We also assessed association of *ACE2* expression with adipose tissue estimated cell type proportions, specifically each of macrovascular endothelial cell, macrophage and adipocyte proportions, estimated as described above.

We performed rank-based inverse normal transformation of all quantitative phenotypic traits. Association between *ACE2* gene expression levels and each phenotypic trait was assessed by fitting linear mixed effects models using the lmer function from the lme4 package^57^ (TwinsUK) or the lm function (METSIM and FUSION) in R^58^ version 3.5.1 (TwinsUK and METSIM) or version 3.6.3 (FUSION). Inverse normalised *ACE2* expression was treated as a continuous dependent variable, and each phenotypic trait in turn was treated as an independent fixed effect.

Covariates included in the TwinsUK study were: age, BMI (for all phenotypes except BMI and obesity), number of hours fasted, sample median transcript integrity number (TIN)^59^, median insert size, and mean GC content included as fixed effects. Random effects included were date of RNA sequencing, morning *versus* afternoon visit time, RNA extraction batch, primer index, family and zygosity. Family and zygosity are both random effects that describe the family a twin belongs to, and their clonality (MZ/DZ status). Each family identifier is specific to a twin pair, and each zygosity label is identical for each pair of MZ twins, while individual DZ co-twins are each given a unique zygosity label. In TwinsUK, the full model was then compared to a null model where the phenotypic trait of interest was omitted, using a 1 degree of freedom ANOVA.

Covariates included in the METSIM study were: age, BMI (for all phenotypes except BMI and obesity), blood cell type proportion, read deletion size, mean read insertion size, TIN, and sequencing batch.

Covariates included in the FUSION study were: age, sex, BMI (for all phenotypes except BMI and obesity), collection site, blood cell type proportion, four genetic PCs, mean GC content, median read insertion size, RNA integrity number (RIN), TIN, and sequencing batch included as fixed effects. In the FUSION study, which included both males and females, analyses were also performed in males and females separately, excluding the sex covariate in the models.

Sensitivity analyses were conducted for all three studies, including estimated MVEC cell type proportion, then both estimated MVEC and macrophage cell type proportions, as an additional covariate(s) in the models above.

### Meta-analysis of phenotypic associations with adipose tissue *ACE2* expression

We conducted a random-effects inverse-variance weighted meta-analysis of up to 1,237 participants, combining *ACE2* gene expression-phenotype association results from the TwinsUK, METSIM and FUSION studies (See Supplementary Tables 1-4 for trait-specific numbers of participants). Meta-analysis was conducted for 13 out of 15 traits using the rma function (restricted maximum likelihood method) from the metafor package^60^ in R version 3.5.1^58^. Association of T2D with *ACE2* was assessed in TwinsUK only, as it was the only study with participants with established T2D at time of biopsy. Association of *ACE2* expression with sex was assessed in FUSION only, given that the TwinsUK and METSIM studies included only females or males, respectively, to avoid biasing of estimates that may result from confounding by study-specific technical differences.

A multiple testing correction threshold of *P* < 3.33 × 10^-3^, calculated using a Bonferroni correction for 15 traits, was used to assess significance of phenotypic associations with *ACE2* expression.

### *ACE2* heritability analyses

Heritability of gene expression levels of *ACE2* was assessed in the TwinsUK sample using an ACE model to decompose variance in gene expression residuals, adjusted for technical covariates, into additive genetic effects (A), common environmental effects shared by both mono-and dizygotic co-twins (C) and unique environmental influences (E). ACE models were fitted using the twinlm function from the mets package^61,62^ in R version 3.5.1^58^. Heritability models included age as a covariate.

### Genotype data, imputation and quality control

In TwinsUK, imputed genotype data and adipose tissue gene expression data were available for 722 females. TwinsUK, METSIM and FUSION genotype data were generated as previously described^63,46,64^, and were imputed to the Haplotype Reference Consortium panel (HRC version 1.1). Variants with MAF < 0.01, imputation R^2^ < 0.5, and Hardy Weinberg *P* < 1 × 10^-6^ were excluded. HRC-imputed genotype data were used for genome-wide association analysis of *ACE2* expression for all studies.

For *ACE2* ciseQTL analyses, in TwinsUK, HRC-imputed genotypes (HRC version 1.1) were available only for autosomes. Chromosome X genotypes were called from low depth 7x sequencing as part of the UK10K project, as previously described^65^ and were available for 490 of 765 TwinsUK participants for whom adipose tissue gene expression data were available. 39 participants were excluded from cis-eQTL analyses due to skewing of X chromosome inactivation^66^. In METSIM and FUSION, HRC-imputed genotypes (HRC version 1.1) were available for all chromosomes including the X in all participants^46,64^. In males, chromosome X variants were coded as 0,1 alleles.

### Cis-eQTL analyses of adipose tissue *ACE2* expression

We defined proximal genetic variants to be those within 1Mb on either side of the *ACE2* transcription start site. For TwinsUK, we adjusted gene counts for family structure using a linear mixed effects model including family and zygosity as random effects. In each study, cis-eQTL analyses were conducted using QTLTools, including BMI and 40 PEER factors^67^ as covariates.

### Genome-wide association analysis of *ACE2* expression

We conducted a GWAS of *ACE2* expression, in order to identify trans-eQTLs that may modulate *ACE2* expression. All variants greater than 1 Mb from the *ACE2* transcription start site were included in this analysis. For TwinsUK, *ACE2* rank inverse normalised gene counts were first adjusted for family structure and technical covariates using a linear mixed effects model including mean GC content and median insert size as fixed effects and sample processing date, primer index, RNA extraction batch, family and zygosity as random effects. GWAS of *ACE2* expression rank inverse normalised expression (METSIM and FUSION) or residuals as described above (TwinsUK) was then conducted using QTLTools^49^ for all studies. Covariates included age, BMI, sample median TIN^59^ and genotyping chip as covariates (TwinsUK). For METSIM, covariates included age, BMI, read deletion size, mean read insertion size, sample median TIN, sequencing batch, and blood proportion as covariates. For FUSION, covariates included were sex, age, BMI, RIN, median TIN, collection site, batch, four genetic PCs, mean GC content, and median insert size as covariates. FUSION also performed sex-stratified GWAS using the same covariates except sex.

### Meta-analysis of *ACE2* expression cis-eQTL and GWAS association results

Cis-eQTL and genome-wide association analyses of adipose tissue *ACE2* expression from the TwinsUK, METSIM and FUSION cohorts were meta-analysed using the sample-size based method implemented in METAL^68^. Meta-analysis was limited to variants present in a minimum of two out of the three studies (cis-eQTL: N_subjects_ = 1,157, N_variants_ = 2,669 variants; *ACE2* expression G^VAS: N_subjects_ = 1,428, N_variants_ = 7,652,879 variants). Due to the fact that the *ACE2* gene is located on the X chromosome, we conducted meta-analyses including all participants, as well as sex-stratified.

The threshold for significance of association of cis-eQTL variants with *ACE2* expression was *P* < 6.94 × 10^-5^, calculated using the EigenMT approach to estimate the number of independent tests in the region in the largest sample (TwinsUK). 721 principal components were found to account for > 99% of the variance in the variants tested. For the identification of trans-eQTL variants associating with *ACE2* expression, a genome-wide threshold of significance of *P* < 5.0 × 10^-8^ was used.

### *ACE2* methylation analyses in TwinsUK

DNA methylation patterns were profiled with the Illumina Infinium HumanMethylation450 BeadChip for 540 adipose tissue biopsy samples, as previously described^35^. Methylation β-values were pre-processed with the ENmix package^69^ within the R environment and inverse-normalised. After filtering out samples skewed for X-chromosome inactivation, there were 495 samples for downstream analyses. For GWAS of DNA methylation levels in CpGs annotated within the *ACE2* gene (i.e. methylation quantitative trait loci, meQTLs) DNA methylation values were first adjusted for covariates in a linear mixed-effects model with fixed effect factors including smoking, age and bisulfite conversion efficiency, and zygosity, and random effects including position on array and family. MeQTL analysis was then performed on the resulting residuals using Matrix eQTL^70^ in both cis (2Mb window, with a total of 325 samples with available genotypes) and in trans for all variants greater than 1Mb either side of the CpGs annotated within the *ACE2* gene. The association between *ACE2* expression and proximal methylation probes was carried out in Matrix eQTL, using methylation and *ACE2* gene expression residuals adjusted for technical and family confounders as previously described (as well as smoking status for methylation data). The methylation-expression association models included age, BMI, T2D status and MVEC estimation as covariates.

### Correlation of phenotypic traits

The correlations between phenotypic traits were assessed in TwinsUK, which was the largest study included in our analyses. Phenotypic trait correlations were assessed using a Spearman correlation in unrelated participants from TwinsUK (n=441), in R version 3.5.1^58^.

### Accession codes

TwinsUK RNA-Seq data are deposited in the European Genome-phenome Archive (EGA) under accession EGAS00001000805. METSIM RNASeq data have been deposited in the Gene Expression Omnibus (GEO) under accession GSE135134. The dbGaP accession number for the FUSION Tissue Biopsy Study is phs001048.v2.p1.

### Code availability statement

All custom scripts, implementing the analyses described in the methods section, are available upon request.

### Display items

## Acknowledgements

KSS acknowledges funding from the Medical Research Council (MR/M004422/1 and MR/R023131/1). TwinsUK is funded by the Wellcome Trust, Medical Research Council, European Union, Chronic Disease Research Foundation (CDRF), Zoe Global Ltd and the National Institute for Health Research (NIHR)-funded BioResource, Clinical Research Facility and Biomedical Research Centre based at Guy’s and St Thomas’ NHS Foundation Trust in partnership with King’s College London. The FUSION and METSIM studies were supported by US National Institutes of Health (NIH) grants U01DK062370, 1-ZIA-HG000024, R01DK093757, R01DK072193, P01HL28481, and Academy of Finland Grants 271961, 272741 and 258753. SMB was supported in part by a grant from the National Institute of General Medical Sciences under award 5T32 GM007092 and acknowledges the Global Partnership Award and UNC Global for travel funds. SV is supported by the National Council of Science and Technology of Mexico (CONACYT).

The authors acknowledge use of the research computing facility at King’s College London, Rosalind (https://rosalind.kcl.ac.uk), which is delivered in partnership with the National Institute for Health Research (NIHR) Biomedical Research Centres at South London & Maudsley and Guy’s & St. Thomas’ NHS Foundation Trusts, and part-funded by capital equipment grants from the Maudsley Charity (award 980) and Guy’s & St. Thomas’ Charity (TR130505). The views expressed are those of the author(s) and not necessarily those of the NHS, the NIHR, King’s College London, or the Department of Health and Social Care. The authors thank Dr. Jörg Saßmannshausen and Dr Lukasz Zalweski from the Rosalind support team for their computational support.

## Author contributions

J.S.E-S.M. and K.S.S. conceived and designed the study. J.S.E-S.M. conducted statistical analyses of the TwinsUK study data and meta-analyses. A.U.J. and L.G. conducted statistical analyses of the FUSION study data. S.M.B. conducted statistical analyses of the METSIM study data. J.S.E-S.M., A.U.J., L.G., S.M.B., L.J.S., K.L.M. and K.S.S. interpreted the data. S.V. performed analysis of the TwinsUK methylation data, supervised by J.T.B. MF advised on statistical analyses. A.L.R., A.Z., L.B., M.R.E., N.N., H.M.S., R.W., T.Y., T.L., S.P., J.T., F.S.C., P.P., M.B., H.A.K. and M.L. participated in sample collection/data generation/data processing. J.S.E-S.M., A.U.J., L.G., S.M.B., A.L.R, L.J.S., K.L.M. and K.S.S. wrote the manuscript. All co-authors provided critical feedback and approved the manuscript.

## Competing interests statement

The authors declare no competing interests.

## Ethics declaration

The TwinsUK project was approved by the ethics committee at St Thomas’ Hospital London, where all the biopsies were carried out. Volunteers gave informed consent and signed an approved consent form prior to the biopsy procedure. Volunteers were supplied with an appropriate detailed information sheet regarding the research project and biopsy procedure by post prior to attending for the biopsy. The FUSION study was approved by the coordinating ethics committee of the Hospital District of Helsinki and Uusimaa, and written informed consent was obtained from all participants. The METSIM study was approved by the Ethics Committee of the University of Eastern Finland and Kuopio University Hospital in Kuopio, Finland, and written informed consent was obtained from all participants.

## Supplementary information

See supplementary information file and supplementary tables. Full summary statistics for meta-analysis of adipose tissue *ACE2* expression GWAS (Supplementary Table 6 of this manuscript) available from the authors upon request.

## Notes

### Competing Interest Statement

The authors have declared no competing interest.

### Funding Statement

Medical Research Council (MR/M004422/1 and MR/R023131/1).
Wellcome Trust
Medical Research Council
European Union
Chronic Disease Research Foundation (CDRF)
Zoe Global Ltd
The National Institute for Health Research (NIHR)-funded BioResource Clinical Research Facility and Biomedical Research Centre based at Guy's and St Thomas' NHS Foundation Trust in partnership with King's College London.
US National Institutes of Health (NIH) grants U01DK062370 / 1-ZIA-HG000024 / R01DK093757 / R01DK072193 / P01HL28481 / and Academy of Finland Grants 271961 / 272741 and 258753.
SMB was supported in part by a grant from the National Institute of General Medical Sciences under award 5T32 GM007092 and acknowledges the Global Partnership Award and UNC Global for travel funds.
SV is supported by the National Council of Science and Technology of Mexico (CONACYT).

